# Genetic architecture of telomere length in 462,675 UK Biobank whole-genome sequences

**DOI:** 10.1101/2023.09.18.23295715

**Authors:** Oliver S. Burren, Ryan S. Dhindsa, Sri V. V. Deevi, Sean Wen, Abhishek Nag, Jonathan Mitchell, Fengyuan Hu, Katherine R. Smith, Neetu Razdan, Henric Olsson, Adam Platt, Dimitrios Vitsios, Qiang Wu, AstraZeneca Genomics Initiative, Veryan Codd, Christopher P Nelson, Nilesh J Samani, Ruth E. March, Sebastian Wasilewski, Keren Carss, Margarete Fabre, Quanli Wang, Menelas N. Pangalos, Slavé Petrovski

## Abstract

Telomeres protect the ends of chromosomes from damage, and genetic regulation of their length is associated with human disease and ageing. We developed a joint telomere length (TL) metric, combining both qPCR and whole genome sequencing (WGS) measurements across 462,675 UK Biobank participants that increased our ability to capture TL heritability by 36% (h^2^_mean=_0.058 to h^2^_combined=_0.079) and improved predictions of age. Exome-wide rare variant (minor allele frequency<0.001) and gene-level collapsing association studies identified 53 variants and 22 genes significantly associated with TL that included allelic series in *ACD* and *RTEL1*. Five of the 31 rare-variant TL associated genes (16%) were also known drivers of clonal haematopoiesis (CH), prompting somatic variant analyses. Stratifying by CH clone size, we uncovered novel gene-specific associations with TL, including lengthened telomeres in individuals with large *SRSF2*-mutant clones, in contrast to the progressive telomere shortening observed with increasing clonal expansions driven by other CH genes. Our findings demonstrate the impact of rare variants on TL with larger effects in genes associated with CH, a precursor of myeloid cancers and several other non-malignant human diseases. Telomere biology is likely to be an important focus for the prevention and treatment of these conditions.

## Introduction

Telomeres are repetitive nucleotide sequences that protect the ends of chromosomes from degradation and are thus considered crucial for maintaining genomic integrity. In somatically dividing cells, telomeres shorten with each replication cycle until they reach a critical length that triggers cellular senescence and ultimately cell death (Rossiello et al. 2022; Harley, Futcher, and Greider 1990). Telomere length (TL) demonstrates considerable interindividual variability and is heritable (Njajou et al. 2007; Broer et al. 2013). Rare germline mutations linked to telomere shortening have been associated with severe diseases, including premature aging syndromes, interstitial lung disease, and immunodeficiencies (Duckworth et al. 2021; Bousfiha et al. 2020; Savage and Alter 2009). Whereas, more subtle reductions in TL have been associated with common, age-related diseases, such as coronary artery disease (Codd et al. 2021). Although TL is heritable, our current understanding of the genetic determinants of TL has been largely limited to the study of common variants. A greater understanding of the genetic determinants of TL could inform disease pathogenesis and expedite the development of novel therapeutic strategies.

High throughput TL assays have been developed to understand telomere biology at the population level. One such method uses quantitative PCR (qPCR) to measure the relative abundance of telomere sequences compared to a reference sequence (Cawthon 2009). More recently introduced *in silico* methods, such as TelSeq, measure average telomere length from whole genome sequencing data (Ding et al. 2014). The advances in genome sequencing of population-scale biobanks provides unprecedented opportunities to leverage these approaches to study the genetic architecture of TL and, ultimately its impact on human health at a population scale. In a recent study of over 400,000 UK Biobank (UKB) participants, a microarray-based genome-wide association study (GWAS) identified over 100 independent common variant loci associated with qPCR TL measurements (Codd et al. 2021). By combining these measurements with whole exome sequencing (WES) data across 418,401 individuals Kessler et al identified rare variant associations for several previously established genes (Kessler et al. 2022). Another study applied the TelSeq algorithm to estimate TL from the whole genome sequences of 109,122 multi-ancestry individuals from the TopMed program and identified thirty-six associated loci, which largely overlap those identified by qPCR based measures (Taub et al. 2022).

Here, we leverage a larger sample size of WGS data from 490,560 multi-ancestry UKB participants to study the genetic architecture of TL, including contributions from both rare and common variants. Moreover, in comparing qPCR- and WGS-derived TL estimates in the same individuals, we observe that combining both measurements into a single statistical metric significantly improves the accuracy of TL estimates and thus empowers discovery potential.

## Results

### Combining qPCR and WGS telomere length estimates increases heritability

Of the 490,560 UKB participants with whole-genome sequencing data, there were 462,675 UK Biobank samples (94%) that met our QC thresholds (Methods) and for whom qPCR TL estimates were available (**Supplementary Table 1** and **Supplementary Fig. 1**). As an orthogonal method for estimating TL, we also used TelSeq, which estimates telomere length from the whole-genome sequencing (WGS) data (Ding et al. 2014).

As expected, TL estimated from TelSeq and qPCR were both significantly associated with age, sex, and ancestry (**Supplementary Fig. 2**). Interestingly, the qPCR- and adjusted TelSeq-TL estimates were only moderately correlated (*r^2^*=0.16; **Fig. 1A**). In a joint model, the association between each of the metrics and age remained highly significant (**Supplementary Table 2**), suggesting that each captures orthogonal information. We derived a PCA linear combination (Aschard et al. 2014) incorporating both qPCR and adjusted TelSeq (**Fig. 1B, Supplementary Figs. 3 & 4**, **Supplementary Table 3**). Using the first principal component, PC1, demonstrated a significant (P < 1 x 10^-16^) performance gain in predicting age compared to models employing either of the individual measures (**Supplementary Fig. 3**).

**Figure 1.**
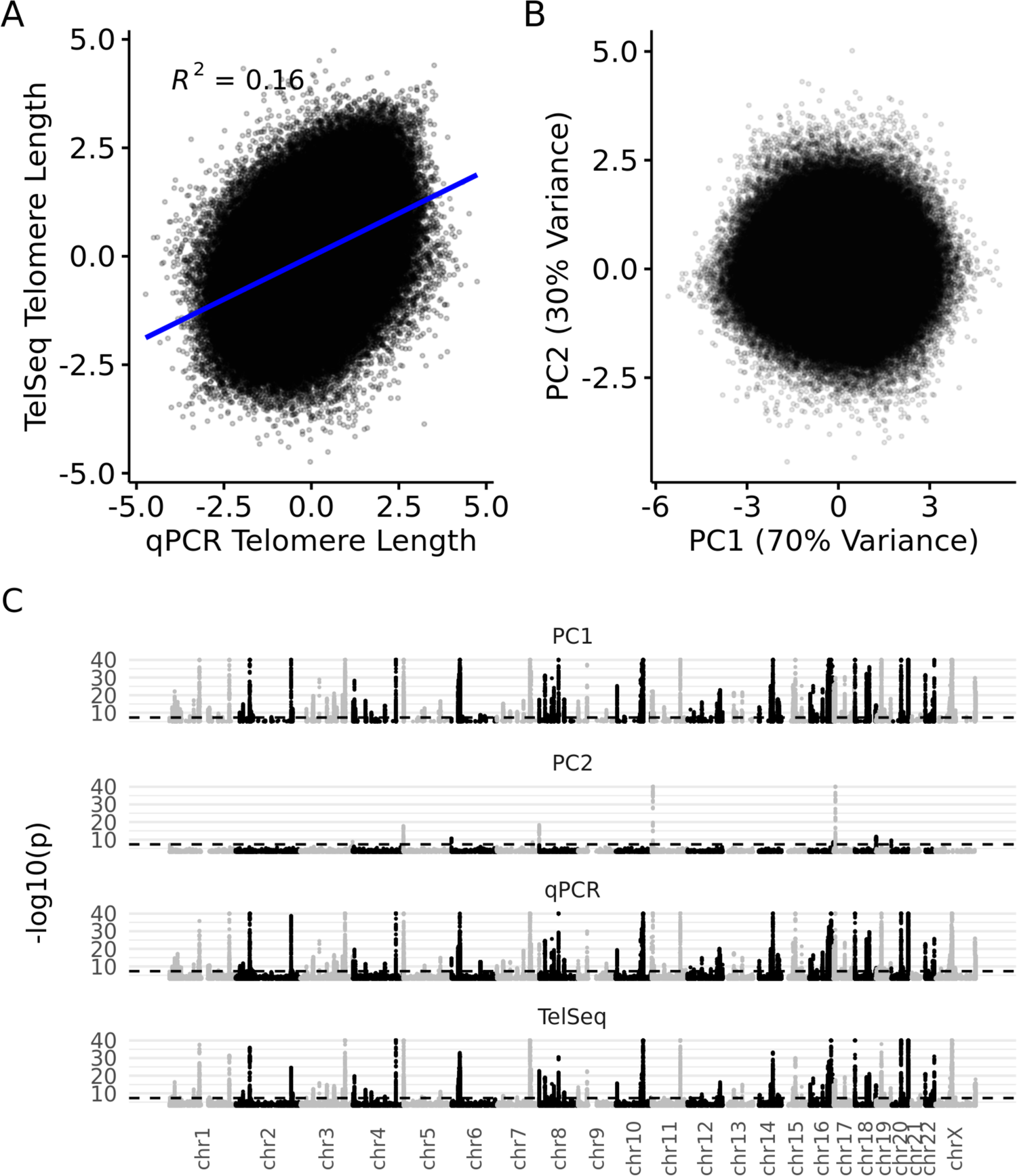
Combining telomere length metrics improves genetic discovery. (**A**) Correlation between inverse normal transformed qPCR and WGS TelSeq telomere length metrics. (**B**) Biplot for PCA analysis of qPCR and TelSeq TL metrics. (**C)** Manhattan plot of common variant analysis of PC1, PC2, qPCR and TelSeq in NFE ancestral group, dotted line indicates P=5 x 10^-8^. For clarity y-axes are truncated at p<1 x 10^-40^.

We first sought to determine common variants (MAF > 0.1%) associated with TL, focusing on 438,359 Non-Finnish European (NFE) ancestry individuals with array-based imputed genotypes available (**Supplementary Table 1**). Using REGENIE (Mbatchou et al. 2021), we performed a common-variant genome-wide association study (GWAS) of TL estimates derived from either qPCR, WGS, PC1, or PC2 (Fig 1C, methods) replicating all signals from *Codd et al.* (**Supplementary Note**). LD-score regression (Bulik-Sullivan et al. 2015) revealed that the PC1 vector had the highest heritability (h^2^=0.079, S.E +/- 0.009, **Supplementary Table, 4**), suggesting the combined TL metric explains more TL variance due to genetic variation than either qPCR or TelSeq alone.

We undertook single variant fine-mapping for all significant (p<5 x 10^-8^) loci (excluding the major histocompatibility region) in the qPCR, TelSeq, and PC1 GWAS. The PC1 TL score resulted in smaller 95% credible SNP sets (median=9) compared with the separate qPCR and WGS GWASs (median=12 and 15, respectively), highlighting that PC1 can more effectively highlight potentially causal variants. In total for PC1 we identified 162 significant (p<5 x 10^-8^) loci (**Supplementary Tables 5, 6**), 39 of which were not within 1Mb of a previously implicated locus. Associations at known loci were also stronger with PC1 compared with qPCR or TelSeq, further demonstrating the value of the combined metric (**Supplementary Figure 5**).

There were also ten significant loci identified in the PC2 GWAS (**Supplementary Tables 5, 6**), most of which were driven exclusively by a single underlying TL metric (**Supplementary Figure 6**). Moreover, 70% of these associations (n=7/10; 3q29:*LMLN*, 5p15.33:*PLEKHG4B*, 6p25.3:*DUSP22*, 7q36.3:*VIPR2*, 16q24.3:*PRDM7*, 18q23:*PARD6G* and 20p13:*DEFB125*) were peri-telomeric (< 2Mb). There was one qPCR association at 11p15.4 (rs1609812) proximal to *HBB* (P=8.3 x 10^-60^ beta=-0.05 [−0.05 to −0.04]), which is used as the reference gene to normalise the qPCR TL assay and has been previously thought to be driven by artefactual technical signals (Codd et al. 2021). Consistent with this being a putative qPCR TL artifact, this locus was not significant in the TelSeq GWAS (P=0.85, beta=0.005 [0.005 to 0.006], **Supplementary Fig. 7**). Collectively, these results demonstrate the superior performance of a linear combination of TL metrics to detect associations and further highlight PC2’s potential to flag spurious associations.

### Rare variant analysis of telomere length reveals allelic heterogeneity

We observe that rare variants have demonstrably larger effects on TL than common variants and have also been implicated in numerous telomere-related diseases. Here, we focused on protein-coding variants observed in whole-genome sequencing data from 439,491 UK Biobank participants of Non-Finnish European (NFE) ancestry to examine the effect of rare variation on PC-derived TL estimates. We performed both variant-level (exome wide association study, ExWAS) and gene-level (rare variant aggregated collapsing analyses) as previously described (Wang et al. 2021). We observed high concordance (*r^2^* = 0.99) between the effect sizes for the common variants included in the ExWAS and our separate common variant GWAS (microarray genotyping) analyses. Genomic inflation was also well-controlled with a median λ_GC=_1.08 (**Supplementary Figure 8**).

We restricted our downstream analyses of the ExWAS to rare (MAF<0.1%) exonic variants that were too rare to be well-represented in the GWAS. Based on our previously identified significance threshold of p≤1}10^-8^(Wang et al. 2021), there were 46 significant rare variant germline associations across 17 distinct genes (**Fig. 2a**, **Supplementary Table 7**) for PC1 after excluding variants that were also significantly associated with PC2 (**Supplementary Figure 9**). Although all of the variants except 9-136496196-CAG-C (*NOTCH1*.p.Pro2514fs P=3.7 x 10^-12^ beta=-2.75 [−3.52 to −1.97]) and 8-84862338-A-G (*RALYL*.p.Ala165Ala P=1.5 x 10^-10^ beta=2.19 [1.52 to 2.86]) overlapped with a previously identified GWAS locus, the absolute effect sizes observed for the ExWAS analyses were generally significantly greater than that previously reported for the same loci. Of the 46 rare variant germline signals, 24% (11/46) were only significantly associated with PC1 and not underlying qPCR or TelSeq measurements.

**Figure 2.**
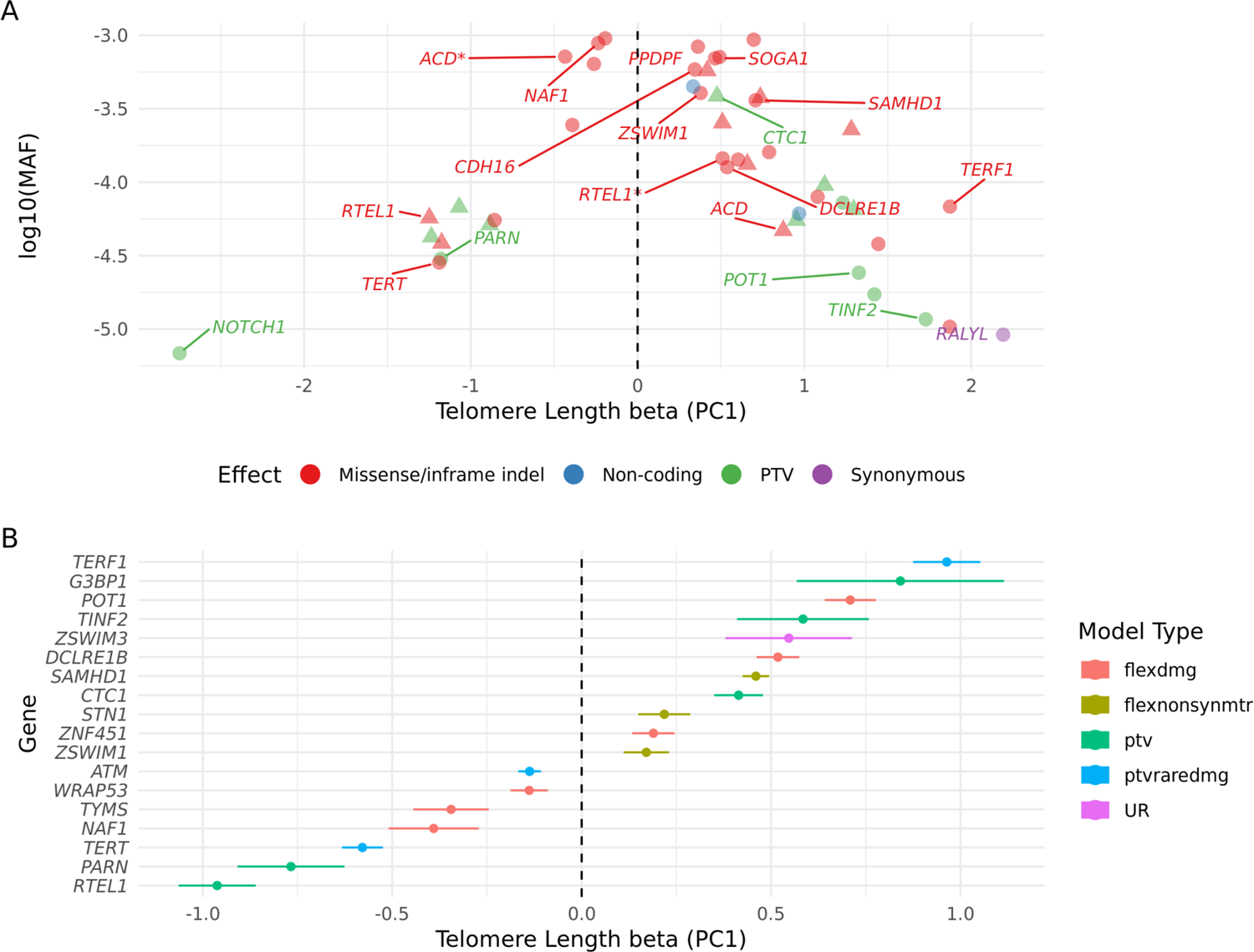
Rare variant analysis of telomere length. **(A)**. ExWAS analysis of PC1 TL, only rare germline variants significant (P ≤ 1}10-8) for PC1 and not PC2 are shown, for clarity the variant with the largest effect for a gene is labelled, variants with opposing effect size in the same gene are starred and triangles indicate HGMD pathogenic variants. **(B)** Collapsing analysis of PC1, the most significant (p<=1}10^-8^) association for a gene over all qualifying variants models (**Supplementary Table 9**) is shown, associations driven by putative somatic variants are excluded.

Thirty-two germline rare variants were associated with longer TL and clustered in components of the CST (*CTC1*) and Shelterin (*ACD*, *TERF1*, *TINF2 POT1*) complexes, both of which function to protect telomere ends and regulate interactions with telomerase. Of these, eight were protein truncating variants (PTVs) in *CTC1*, *POT1, TINF2,* and *TERF1*, all of which are genes implicated in telomere-associated diseases. Interestingly the PTV in *CTC1* (17-8237439-GCTTT-G p.Lys242fs P=2.3 x 10^-19^ beta=0.48 [0.37 to 0.58]) has been implicated in compound heterozygous recessive cerebroretinal microangiopathy with calcifications and cysts (CMCC, also known as Coats plus syndrome), which is associated with shorter telomeres (Anderson et al. 2012; Gu and Chang 2013). Our results indicate that outside of the context of nullizygosity this PTV is associated with longer TL, concordant with prior observations of CTC1 depletion promoting excessive telomerase activity (L.-Y. Chen, Redon, and Lingner 2012). We also observed three PTVs associated with TL in *POT1*, which is associated with Familial Glioma, Familial Melanoma, cardiac angiosarcoma and chronic lymphocytic leukaemia (CLL) (DeBoy et al. 2023; Bainbridge et al. 2015; Speedy et al. 2016; Calvete et al. 2015; Shi et al. 2014).

Remarkably, the remaining 14 rare non-synonymous germline variants associated with shorter TL and were clustered in genes previously associated with autosomal dominant Dyskeratosis Congenita and/or pulmonary fibrosis (*ACD*[OMIM:609377]*, PARN*[OMIM:604212]*, RTEL1*[OMIM:608833]*, NAF1*[OMIM:620365] and *TERT*[OMIM:613989]). In both *ACD and RTEL1,* we observed independent rare non-synonymous variants with opposing effects indicating a possible allelic series in these two genes. For example, in *ACD* two rare missense variants clustering within the *POT1* binding domain (16-67659046-C-A p.Arg259Leu and 16-67659234-T-C p.Asn246Ser) were associated with increased TL and one (16-67660036-C-T p.Asp120Asn) in the N-terminal oligonucleotide/oligosaccharide-binding (OB) domain that acted in the opposite direction (**Table 1**). *ACD* encodes TPP1, a key component of the 6 protein shelterin complex. Consistent with our results, a recent mutagenesis revealed that mutations that disrupt POT1 binding promote ectopic initiation of ATR- and ATM-mediated DNA damage repair programs, resulting in longer telomeres (Grill et al. 2021). Reciprocally, mutations within the N-terminal OB are associated with disrupted telomerase recruitment leading to progressively shorter TL (Grill et al. 2021), mirroring the effect of the 16-67660036-C-T variant we detected in this region.

**Table 1.** Rare variants in *ACD* modulating telomere length. *. Protein coordinates with respect to Uniprot (Q96AP0) canonical transcript ENST00000620761.6. **Also detected through our GWAS.

| RS Number | Variant ID | MAF | Effect | P-value | Consequence* | Domain |
| --- | --- | --- | --- | --- | --- | --- |
| **rs139438549\*\*** | 16-67658960-T-C | 0.001 | 0.43 | $9.9 \times 10^{-11}$ | Thr205Ala | POT1 binding domain |
| **rs145007645** | 16-67659046-C-A | $1.6 \times 10^{-4}$ | 0.74 (0.63 to 0.95) | $2.0 \times 10^{-22}$ | Arg176Leu | |
| **rs370512338** | 16-67659234-T-C | $3.7 \times 10^{-4}$ | 0.74(0.63 to 0.84) | $2.3 \times 10^{-44}$ | Asn163Ser | |
| **rs249052024** | 16-67659240-G-A | $6.4 \times 10^{-4}$ | -0.26 (-0.34 to -0.18) | $1.41 \times 10^{-10}$ | Ser161Leu | |
| **rs142662151** | 16-67660036-C-T | $7.2 \times 10^{-4}$ | -0.43 (-0.51 to -0.40) | $3.2 \times 10^{-12}$ | Asp37Asn | OB1 |

Although less frequent than common variants, rare variants can still be correlated due to linkage disequilibrium (LD). To resolve signal independence among the rare variants, we performed conditional analyses (methods) and found that one of our signals: *SOGA1* (20-36810011-C-T p.Ala852Thr P=1.9 x 10^-32^ beta=0.46 [0.38 to 0.54]) is likely due to LD with a *SAMHD1* 20-36898455-C-G signal (**Supplementary Table 8**). *SOGA1* is thus unlikely to constitute a novel TL related gene.

### Rare variant gene-level collapsing analysis

We performed gene-level collapsing analyses to identify genes associated with telomere length through the aggregated presence of variants too rare and thus underpowered to be individually discovered in ExWAS analyses. As previously described, we employed ten-qualifying variant (QV) models (Wang et al. 2021)(**Supplementary Table 9**), and association statistics were well-calibrated with a median λ_GC=_1.07 (**Supplementary Fig. 10**). After filtering putative somatic signals we identified 18 genes significantly (p≤1}10^-8^) associated with PC1 TL, 3 (17%) of which were uniquely identified in PC1 and not the individual qPCR or TelSeq statistics (**Fig. 2b**, **Supplementary Table 10**, **Supplementary Fig. 11**).

Fourteen of the gene-level signals arose from the rare protein-truncating “PTV” QV model. Five of these genes were associated with telomere shortening (*ATM*, *RTEL1, PARN, TERT* and *NAF1*) and all five have been implicated in known telomere-related clinical diseases, including pulmonary fibrosis (IPF) (Stanley et al. 2016; Stuart et al. 2015; Dhindsa et al. 2021) and Dyskeratosis congenita (Revy, Kannengiesser, and Bertuch 2023). The remaining nine PTV collapsing model signals associated with longer TL. Seven of these nine genes have established biological roles in protection from TL attrition (*POT1*, *TERF1, TFIN2*, *CTC1* and *STN1*), DNA-repair (*DCLRE1B;* formerly *APOLLO*), and thymidine nucleotide metabolism (*SAMHD1*) (Mannherz and Agarwal 2023)).

Two genes significantly associated with longer TLs in the rare PTV collapsing model have not been previously described in increased telomere length biology. *G3BP1* (P=1.2 x 10^-9^ beta=0.84 [0.57 to 1.11]), encodes an RNA-binding protein involved in RNA metabolism regulation and stress granule formation.(Ge et al. 2022) It is also known to bind guanine quadruplexes (G quadruplexes), which are a substrate for human telomerase(Bryan 2020; Moye et al. 2015). The other gene, *ZNF451* (P=8.1 x 10^-9^ beta=0.30 [0.20 to 0.41]), encodes a Zinc finger protein that acts as a SUMO ligase and a DNA repair factor that controls cellular responses to TOP2 damage (Park et al. 2023).

There were several other novel significant associations that arose in the QV models that included protein-truncating variant effects alongside putatively damaging missense variants. *TYMS* (flexdmg P=3.1 x 10^-12^ beta=-0.34 [−0.44 to −0.25]), which has also been observed as a hit in a CRISPR-Cas9 screen for telomere length (Mannherz and Agarwal 2023) and has been causally associated with Dyskeratosis congenita (Tummala et al. 2022), was associated with reduced telomere length. *WRAP53* (flexdmg P=5.9 x 10^-9^ beta=-0.14 [−0.19 to −0.09]), which encodes a component of the telomerase holoenzyme complex, was also associated with decreased telomere length. The *ZSWIM1* (UR, P=5.0 x 10^-9,^ beta=0.17 [0.11 to 0.23]) and *ZSWIM3* (flexnonsynmtr, P=7.4 x 10^-11^, beta=0.55 [0.38 to 0.71]) zinc finger proteins were associated with increased telomere length*. ZSWIM1,* which was also an ExWAS hit, and *ZSWIM3* are in proximity with one another, sitting within a peri-telomeric GWAS locus. We thus performed a leave-one-out analysis (methods), which showed that no individual variants in *ZWIM1* and/or *ZSWIM3* were responsible for driving either gene-level association (**Supplementary Fig. 12**). Moreover, conditional analysis indicated that both *ZSWIM1* and *ZSWIM3* associations were independent of each other and of the 20-45884012-G-A *ZSWIM1* missense variant identified from our ExWAS analysis. Altogether, the rare-variant aggregated gene-level collapsing analysis framework uncovered several loci that were not detectable in the variant-level analyses.

### Multi-ancestry rare-variant analysis

Including individuals of non-European ancestries is critical for health equity and bolstering gene discovery (Petrovski and Goldstein 2016; Ben-Eghan et al. 2020). Therefore, we performed additional GWAS, ExWAS, and collapsing analysis on PC1 in five additional UK Biobank ancestral groups (AMR, EAS, SAS, ASJ and AFR; **Supplementary Table 1**). The ancestry GWAS revealed a single locus in the AFR ancestry cohort that was not detected in the NFE analyses (rs146660284, P_AFR=_2.5 x 10^-8^, beta_AFR=_0.67 [0.43 to 0.90]) and there were no non-NFE ancestry-specific rare variant associations, likely due to the substantially smaller sample sizes of these populations in the UK Biobank. A fixed effect meta-analyses was then performed to combine results across ancestral strata, which detected an additional 4 loci (**Supplementary Table 11**) through the GWAS and one further rare protein-coding variant missense association in *RTEL1* (20-63692865-C-G p.Gln682Glu P=7.4 x 10^-9^ beta=-0.77 [−1.03 to −0.51]). For the collapsing meta-analysis, no new study-wide significant genes were identified; however, there was a consistent improvement in observed statistical power indicating that future cross ancestry sequencing studies are likely to identify further causal gene TL associations (**Supplementary Fig. 13**).

### Association between telomere length and clonal haematopoiesis

Telomere length has been shown to be causally associated with clonal haematopoiesis (CH) (Nakao et al. 2022). In our rare variant analyses, we identified several TL associations with five known CH driver genes (ExWAS: *CALR* and *JAK2*, Collapsing: *CALR*, *TET2*, *ASXL1*, and *PPM1D*) (**Supplementary Tables 7 and 10**), which we reasoned are likely driven by somatic events rather than germline inherited variation (**Supplementary Fig. 14**). To investigate this further, we performed somatic variant calling in 15 established CH and myeloid cancer driver genes (**Supplementary Table 12**) using the complementary UK Biobank higher coverage exome sequencing data (Dhindsa et al. 2022). Using these somatic CH calls, and adjusting for age, sex and smoking status, we performed collapsing analyses with our PC1 metric and replicated the previously described association between overall CH and shorter TL (Nakao et al. 2022) (**Figure 3A**). Analysing CH driver genes individually, we found that most followed the same pattern of association with shorter TL, including novel observations for *SF3B1* (P=5.8 x 10^-12^, beta=-0.46 [−0.59 to −0.33]) and *PRPF8* (P=0.0028, beta=-0.40 [−0.66 to −0.14]). Conversely, we discovered that CH driven by mutations in *DNMT3A* was significantly associated with longer TL (P=5.81 x 10^-14^, beta=0.07 [0.05 to 0.08]) (**Figure 3A, Supplementary Table 13**).

**Figure 3.**
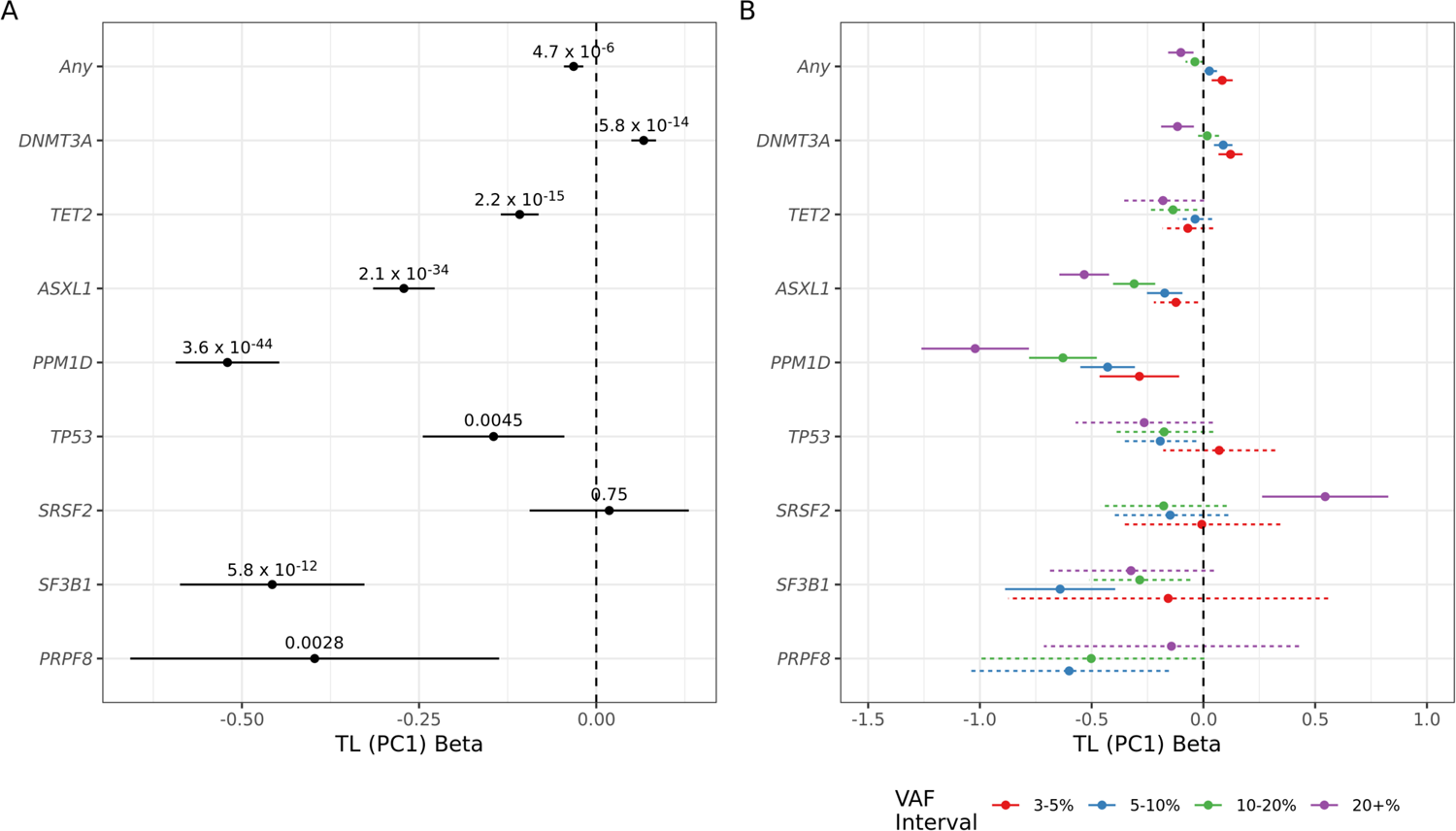
**(A)** Collapsing analysis of somatic variants in select CH genes with TL PC1 horizontal bars indicate 95% confidence intervals and are labelled with P-values. **(B)** Collapsing analysis of somatic variants in CH genes stratified by VAF intervals (colours), associations not reaching significance are shown with dashed horizontal 95% CI bars. ‘Any’ indicates an overall analysis of the selected CH genes.

To investigate these associations further, and particularly to distinguish cause from effect in the context of TL measures ascertained from bulk blood, we performed subsequent analyses stratifying by the size of the mutant CH clone (**Supplementary Table 12**). Specifically, we reasoned that in individuals with small CH clones (eg. Variant Allele Fraction (VAF)<5%), most blood leukocytes would derive from wild-type (non-CH) cells and therefore reflect background TL. In comparison, in individuals with larger CH clones, average TL across blood cells would increasingly reflect TL within the mutant CH clone itself.

Small clones (e.g. VAF 3 – 5 %) were associated with longer TL for overall CH (P=4.8 x 10^-4^, beta=0.08 [0.04 to 0.13]) and *DNMT3A*-mutant CH (P=1.0 x 10^-5^, beta=0.12 [0.07 to 0.18), consistent with previous reports that longer TL promotes CH acquisition (**Figure 3B, Supplementary Table 14**) (Nakao et al. 2022; DeBoy et al. 2023). However, intriguingly, we discovered the inverse association for some other CH drivers, where small clones were associated with shorter TL, suggesting that acquisition of certain CH subtypes are promoted by shorter telomeres. A notable example was *PPM1D*, consistent with reports of high prevalence of *PPM1D*-mutant CH in individuals with inherited short telomere disorders (Ferrer, Mangaonkar, and Patnaik 2022).

Also aligning with previous reports, for CH overall and for most individual CH driver genes, we observed progressive shortening of TL with increasing clone size (Any P=1.1 x 10^-11^ beta=-0.42 [−0.54 to −0.30]), likely reflecting accelerated telomere attrition with cell division in expanding clones (**Supplementary Table 15**). However, a striking exception to this pattern was observed in *SRSF2*-mutant CH, in which large clones were unexpectedly associated with longer TL (P=5.37 x 10^-6^ beta=1.34 [0.77 to 1.90]), suggesting that *SRSF2* mutations may mediate telomere elongation in CH.

## Discussion

This study of 462,675 multi-ancestry individuals presents the most extensive genetic interrogation of TL to date. Importantly, we discovered that qPCR- and WGS-derived estimates of TL capture orthogonal data. Combining these metrics via PCA not only enhanced downstream analyses, but also allowed us to discriminate artefactual signals (i.e., associations with PC2). This has important implications for future population-based studies, as it suggests that, where possible, the most robust assessments should leverage both metrics.

Through both common and rare variant-oriented studies, we described several novel TL loci that give insight into telomere biology. For example, we uncovered antagonistic allelic heterogeneity in *ACD* and *RTEL*1, highlighting the complex role for rare variants in telomere homeostasis and their role in disease. Moreover, the disease associations with both shorter and longer TL underscores the challenge of therapy development, where perturbation of balanced antagonistic effects might lead to significant off-target effects. We also identified a previously undescribed association between PTVs in *G3BP1* and longer TL. While *G3BP1* is involved in stress granule formation, its role in mediating TL is currently unclear and will require functional work in future studies.

Previous studies (Kessler et al. 2022; Nakao et al. 2022) have highlighted a causal, bi-directional relationship between TL and CH. Here, we uncovered novel driver gene-specific links between CH and TL, providing new insights into the mechanisms driving clonal expansion. Longer telomeres predispose to *DNMT3A*-mutant CH, perhaps by extending cellular replicative potential, whereas this is not the case for some other CH driver genes, including *PPM1D*. It is notable that *PPM1D*-mutant CH is known to be particularly enriched among individuals with inherited short telomere disorders(Ferrer, Mangaonkar, and Patnaik 2022) and in individuals exposed to DNA-damaging chemotherapies that appear to shorten telomeres(Ishibashi and Lippard 1998; Saker et al. 2018; Kahn et al. 2018). Taken together, we hypothesise that *PPM1D* mutations are specifically advantageous to blood stem cells in the context of critically short telomeres, perhaps by conferring resistance to the replicative senescence that would ordinarily occur in this setting.

It is also notable that mutations in particular splicing genes, such as *SRSF2*, have been shown to drive CH exclusively in older individuals (Fabre et al. 2022), by which time telomeres have naturally shortened with age. The discovery that telomeres in *SRSF2*-mutant CH do not appear to shorten as clones expand, or even to elongate, contrasts starkly with the accelerated attrition of telomeres with clonal expansion driven by other CH genes. The possibility that *SRSF2* mutations confer advantage through telomere modulation offers a novel explanation for the expansion of these mutant clones specifically in older age. In summary, our findings support a key role for telomere maintenance in the development of CH, via mechanisms specific to the mutant gene driving clonal expansion. Since CH is a causal risk factor for progression to myeloid cancers and for a range of non-haematologic diseases, with larger CH clones conferring higher risks (Weeks et al. 2023; Jaiswal 2020), therapeutic modulation of telomere biology might be an important focus as strategies for prevention and treatment of CH and its sequelae.

## Methods

### Cohort description

Whole genome sequences (WGS) were available for 490,560 UK Biobank participants. 490,503 (99.99%) of sequences remained after removing contaminated sequences (verifybamid_freemix >= 0.04) using VerifyBAMID (Jun et al. 2012) or that had low CCDS coverage (<94.5% of CCDS r22 bases covered with ≥10-fold coverage). A further 106 sequences were removed after being identified as sample duplicates with multiple birth events. For the remaining 490,397 WGS we used KING(Manichaikul et al. 2010) to identify individuals with first-degree relatives, which we then randomly pruned such there were no pairs of samples with a kinship coefficient > 0.354 to leave 490,216 (99.93%) WGS. We used peddy(Pedersen and Quinlan 2017) and 1000genomes data to classify ancestries (peddy_prob>=0.9) using the gnomAD classifier(S. Chen et al. 2022) to subdivide EUR into individuals of non-Finnish (NFE) and Ashkenazi Jewish (ASJ) ancestries. We performed additional QC on NFE ancestry samples using peddy-derived principal components (PC) removing samples that fell outside of 4 standard deviations from the mean over the first four PCs. Finally, we removed sex-discordant samples to leave 482,848 (98.4%) of samples for analysis. Final cohort sizes stratified by ancestry are indicated in **Supplementary Table 1**.

### Whole Genome Sequencing processing and variant calling

Whole-genome sequencing (WGS) data of the UKB participants were generated by deCODE Genetics and the Wellcome Trust Sanger Institute as part of a public-private partnership involving AstraZeneca, Amgen, GlaxoSmithKline, Johnson & Johnson, Wellcome Trust Sanger, UK Research and Innovation, and the UKB. These individuals were pseudo randomly selected from the set of UKB participants. The WGS sequencing methods have been previously described (Halldorsson et al. 2022). Briefly, genomic DNA underwent paired-end sequencing on Illumina NovaSeq6000 instruments with a read length of 2×151 and an average coverage of 32.5x. Conversion of sequencing data in BCL format to FASTQ format and the assignments of paired-end sequence reads to samples were based on 10-base barcodes, using bcl2fastq v2.19.0. Initial quality control was performed by deCODE and Wellcome Sanger, which included sex discordance, contamination, unresolved duplicate sequences, and discordance with microarray genotyping data checks.

UK Biobank genomes were processed at AstraZeneca using the provided CRAM format files. A custom-built Amazon Web Services (AWS) cloud compute platform running Illumina DRAGEN Bio-IT Platform Germline Pipeline v3.7.8 was used to align the reads to the GRCh38 genome reference and to call small variants, including on the mitochondrial genome where a continuous allele frequency model is used; a single alternate allele is considered as a candidate variant and an allele fraction is estimated for emitted variants. All PASS variants emitted had a confidence score (LOD) above the default of 6.3. Variants were annotated using SnpEff v4.3(Cingolani et al. 2012) against Ensembl Build 38.92(Zerbino et al. 2018).

### Whole-Exome Sequencing

Full details of the whole exome sequencing and subsequent variant calling and annotation of the UKB cohort are described fully in Wang et al (Wang et al. 2021). Briefly, genomic DNA underwent paired-end 75-bp whole-exome sequencing at Regeneron Pharmaceuticals using the IDT xGen v1 capture kit on the NovaSeq6000 platform. Reads were aligned to GRCh38 and small indels and SNVs called using running Illumina DRAGEN Bio-IT Plat-form Germline Pipeline v3.0.7. The resultant catalogue of variants was annotated using snpEFF v4.3(Cingolani et al. 2012), Ensembl v38.92(Zerbino et al. 2018), REVEL(Ioannidis et al. 2016), and MTR(Traynelis et al. 2017) scores.

### Estimating telomere length from WGS data

We used TelSeq (Ding et al. 2014) v0.0.2 to estimate telomere length using whole genome sequencing data in 482,848 UKB individuals. We used readlength (-r) 150 and kmer size (-k) 10 to match the proportion threshold (40%) for a read to be classified as of telomeric origin as described in Ding et al.

### Correlation analysis

In total 462,675 samples had TL estimates from both TelSeq and qPCR methods and pairwise Pearson correlation was assessed using the R ‘cor’ function. To assess the contribution and degree of collinearity between Telseq and qPCR methods we fit the following model linear model using inverse rank normal transformed age, TelSeq and qPCR (adjusted T/S ratio - UKB field 22191)

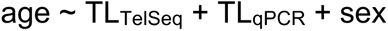

We then used the R package olsrr (v0.5.3) to compute variance inflation factors (VIF) for each of the predictors, finding a mean VIF of 1.125 indicating no evidence of collinearity. Overall removing TL_TelSeq o_r TL_qPCR f_rom the model reduced R^2^ by 0.10 and 0.14 respectively.

### WGS TL measurement confounder analysis and adjustment

We fit a linear model to NFE ancestry samples, using TelSeq, qPCR TL metrics as dependent variables selecting the available (and non-colinear) WGS metrics ‘total read count’, ‘uniformity of coverage’, calling pipeline (deCODE or WTSI), as well as age and sex as a biological control. We used the inverse rank normal transform to scale all variables to facilitate comparison.

We found that that all three WGS metrics were significantly associated with TelSeq TL measurements (**Supplementary Table 3**) and so to adjust for this we refit the linear model, excluding age and sex and taking the residuals as the adjusted TelSeq TL for downstream analyses.

### PCA TL score

Across all 461,461 individuals with both TL measurements, we used the R built-in function ‘*prcomp’* to combine the adjusted TelSeq and adjusted T/S Ratio qPCR(UKB Field 22191) inverse normal transformed TL estimates. Each PCA consisted of two orthogonal principal axes whose sample scores were considered separate TL measurements or ‘TL scores’ with PC1 and PC2 explaining 70% and 30% of the variance respectively.

To assess performance for single and combined TL metrics we randomly sampled 10,000 participants from the full dataset. We used this training set to fit a simple linear model of a given TL metric with age (i.e. age ∼ TL_metric)_. Then using the held-out participants we used the model to predict age and assessed prediction performance as the root mean squared error (RMSE) of the age predictions. To perform cross validation and obtain confidence intervals for these performance estimates we performed this procedure 100 times sampling with replacement.

### NFE GWAS

We used UKB-imputed genotypes (UKB Field 22828) to perform GWAS for qPCR, WGS, qPCR+WGS PC1 and qPCR+WGS PC2. Briefly, we performed additional QC only taking forward NFE samples with imputed genotypes (INFO>0.7, MAC>5) for which all TL metrics were available (n=438,359) We used REGENIE (v3.1)(Mbatchou et al. 2021) with additional covariates of age, sex, genotyping plate, ancestry PCs 1-10 (as supplied by UKB) and WGS sequencing site. We excluded results for SNPs with the following (0.99 missingness, imputation INFO<0.7, and p.HWE > 1 x 10-5). We found no evidence of genomic inflation **(Supplementary Table 4**). We selected sentinel SNPs and EUR-only ancestry summary statistics from Codd et al. for comparison (**Supplementary Figure 5**).

### LD Score regression

We used ldsc (v1.0.1)(Bulik-Sullivan et al. 2015) to assess heritability and further assess possible stratification for each GWAS. Briefly, we used munge_stats.py on the cleaned summary stats (SNPs removed 0.95 missingness, imputation INFO<0.4 and p.HWE > 1e-5), then used ldsc.py to estimate h2 using the supplied 1KG Genomes LD score matrices.

### Defining GWAS loci

To define loci for each phenotype we selected significant variants (p < 5 x 10^-8^), and created regions +/- 1Mb, creating a bespoke region (chr6: 25,500,000 to 34,000,000) for HLA. We then merged overlapping regions by phenotype, for each resultant region, where the most significant variant was selected as the index, in the case of ties the variant closest to the middle of the region was selected. Finally we used the GenomicRanges(Lawrence et al. 2013) *‘reduce’* function to combine overlapping regions regardless of phenotype to define a set of non-redundant loci.

We used GCTA-COJO(Yang et al. 2012) to perform stepwise model selection to define conditionally independent signals for each autosomal locus. Briefly, for each GWAS we selected summary statistics for all variants (INFO>=0.7) where P<1 x 10-6. We then randomly sampled 50,000 individuals from the NFE ancestry cohort for as the LD reference using BGENIX and QUTILS (Band and Marchini 2018) to create bgen files for these individuals. Finally we used PLINK2(Chang et al. 2015) to convert the resultant bgen files to binary PLINK 1.x format suitable for input into GCTA-COJO (gcta 1.94.1 -- cojo-slct) using default settings (--cojo-wind 10000; --cojo-p 5e-8; --cojo-collinear 0.9). For variants on the X chromosome we applied a similar approach but replaced 50,000 reference individuals with 50,000 randomly sampled female individuals of NFE ancestry and due to increased linkage disequilibrium increased window size to 50Mb(Sidorenko et al. 2019).

To assess novelty we compiled a list of significant (p<5e-8) variants from Codd et al.(Codd et al. 2021), Kessler et al.(Kessler et al. 2022), Taub et al.(Taub et al. 2022) and the GWAS catalogue (Sollis et al. 2023) using ‘Telomere Length’ term (EFO_0004505), downloaded on 11/07/2023. We then defined 2Mb regions centred on each variant, and conservatively defined a locus from our study novel if there was no overlap.

### Single causal variant fine mapping

For variant fine-mapping under the single causal variant we selected autosomal variants from NFE GWAS and divided these into approximately independent LD blocks using regions defined in (Berisa and Pickrell 2016). We then used the single variant fine-mapping (Wakefield 2007; Wellcome Trust Case Control Consortium et al. 2012) approach as implemented in https://github.com/ollyburren/rCOGS to assign 95% credible sets.

### ExWAS

We carried out a virtual exome-wide association analysis (ExWAS) of TL using WGS genotypes stratified by NFE (n=439,491), SAS (n=9,349), AFR (n=8,162), EAS (n=2,362), ASJ (n=1,201), and AMR (675) ancestral groups. Briefly we selected unrelated individuals within each ancestry strata with TL and WGS data using the same method as described in ‘Sample QC’. We took forward variants that passed the variant QC as described in Wang et al. which had a MAC>5. We used a linear model of the form TL_PC1∼_ genotype + age + sex + age2 + Peddy_PC1:4 +_ SequenceSite to assess the association of genotype with TL using the R ‘PEACOK’ package(Wang et al. 2021). Here genotype was coded as either a genotypic (AA=0, AB=1, BB=2), dominant (AA=0, AB=1, BB=1) or recessive model (AA=0, AB=0, BB=1) where A and B are the reference and alternate alleles. For NFE ancestral group we assessed 326,846, 326,846 and 62,716 variants for the dominant, genotypic and recessive models respectively (carrier count >=5). For the NFE analyses we report the most significant model-variant pair such that variants P ≤ 1}10^-8^ for PC1 and P >1 } 10^-8^ for PC2 and MAF < 0.1%. For PC1 associated variants passing QC we reran associations analyses for each variant conditional on other significant rare variants within a 2Mb to check for independence.

### Collapsing Analysis

To assess the contribution of very rare variants we carried out a collapsing burden analysis stratified by ancestral groups as per ExWAS analysis, using the method described in Wang et al. Briefly, we aggregated qualifying variants based within the unit of a gene for each ancestral grouping and use these counts in a linear regression using the R ‘PEACOK’ package using the same covariates as for the ExWAS. We defined 10 qualifying variant tests (ST8) that includes a synonymous model as an empirical control. We used the empirical modelling of the null distribution from Wang et al. to define a genome-wide significant threshold of p<1e-8. In total we assessed 18,930 genes across all 10 models. For NFE analyses we report best QV model-gene pair for which P ≤ 1}10^-f^or PC1 and P >1 x 10^-8^ for PC2.

To assess the leverage of individual variants on collapsing analysis genome-wide significant hits we employed a leave-one-out analysis (LOO). For each gene, and qualifying variant model, we reperformed collapsing analysis, leaving out one variant at a time. In this approach variants with a large influence on the overall collapsing analysis, when excluded, result in a concomitant change in statistical significance (**Supplementary Figure 12**).

### Multi-ancestry meta-analysis

We performed inverse variance weighted (IVW) meta-analysis for ExWAS and collapsing across NFE, SAS, AFR, EAS, ASJ, and AMR ancestral groupings for variants with at carrier count >=5 within each grouping. In the context of rare variants IVW can be unstable so we compared IVW meta-analysis P-values with those generated from Stouffer’s method weighting each study by the square root of the sample size. We found that both approaches generated similar p-values indicating that IVW in this setting was stable even for rare variants.

For GWAS multi-ancestral analysis we used REGENIE using the approach described for NFE to generate GWAS summary statistics for SAS, AFR, EAS and AMR samples. We used the locus definition approach described earlier to define significant loci for each ancestral strata, defining novelty as before, considering the PC1 NFE ancestry TL loci previously described. For GWAS we used METAL (Willer, Li, and Abecasis 2010) to perform IVW meta analyses across all ancestry strata. We selected significant variants (P_meta<_ 5 x 10^-8)^ removing those that were present in a single ancestry, using these to define loci and index variants as previously described. We assessed these for overlap with NFE loci defining novelty as before.

### CH Analysis

To detect putative clonal haematopoiesis, we used the pipeline described in Dhindsa et al.(Dhindsa et al. 2022). Briefly, using the same GRCh38 genome reference aligned reads as for WES germline variant calling, we ran somatic variant calling with GATK’s Mutect2 (v.4.2.2.0), After QC we focussed on a set of 15 genes (**Supplementary Table 12**) exhibiting age dependent prevalence for further analyses including only PASS variant calls with 0.03 ≤ Variant Allele Frequency (VAF) ≤ 0.4 and Allelic Depth (AD) ≥ 3 across an annotated set of variants.

For the analysis, we considered four different variant allele frequency (VAF) cut-offs (3-5%, >5-10%, >10-20% and >20%, **Supplementary Table 12**) across NFE ancestry individuals. In total after excluding 3,585 individuals diagnosed with either a haematological malignancy pre-dating sample collection or with a lymphocyte count > 5 x10^9^ cells/litre we took forward 435,525 individuals for analysis. For overall CH driver subtype association (as shown in **Fig1A**) We fit a linear model TL_PC1 ∼_ CH_VAF>0.03 +_ age + sex + age:sex + age2 + ancestry PC1:4 + ever.smoked + pack.years. Where TL_PC1 r_epresents the PC1 telomere length estimate and CH the carrier status for a particular CH driver subtype with VAF> 3%. We then repeated this analysis stratifying by non-overlapping VAF cutoffs for each CH driver subtype. Finally, to get an overall association statistic between TL and VAF stratified by CH driver subtype we repeated this analysis recoding each CH driver gene carrier status by VAF as an ordinal variable.

## Data Availability

All data produced in the present study are available upon reasonable request to the authors

## Notes

### Competing Interest Statement

OB,RD,SV,SW,AN,JM,FH,KS,NR,HO,AP,DV,QW,RM,SW,KC,MF,QW,MP and SP are current employees and/or stockholders of AstraZeneca

### Funding Statement

This study did not receive any funding

### Author Declarations

UK Biobank data use (Project Application Numbers 68601 and 26041) was approved by the UK Biobank according to their established access procedures. UK Biobank has approval from the North West Multi-centre Research Ethics Committee (MREC) as a Research Tissue Bank (RTB), and as such researchers using UK Biobank data do not require separate ethical clearance and can operate under the RTB approval.

### Summary of Updates

Incorrect author name for Ryan Dhindsa

