## Supplementary Information for "Genetic architecture of telomere length in 462,675 UK Biobank whole-genome sequences"

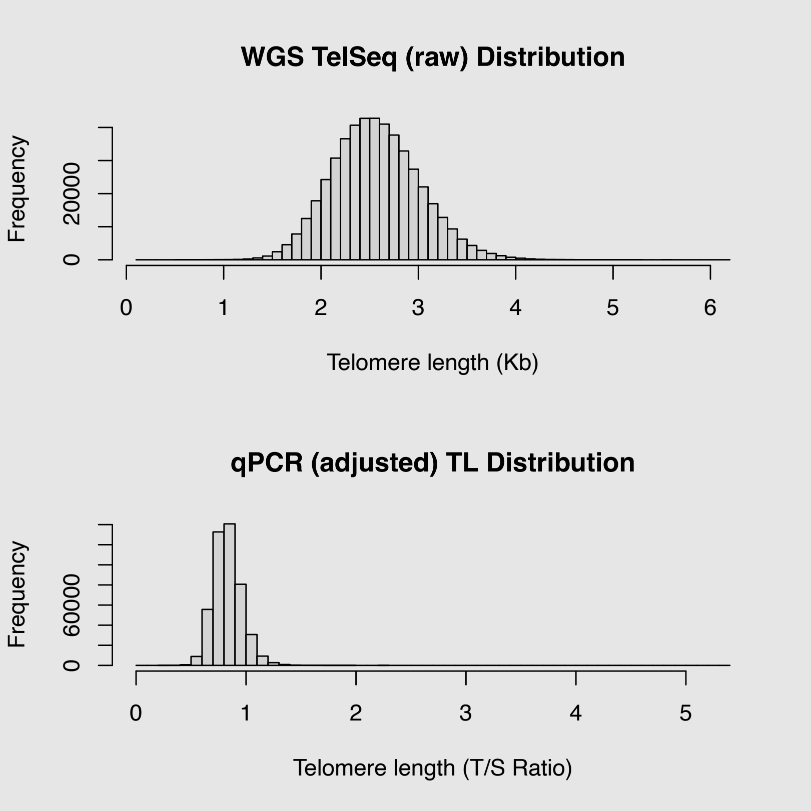


**Supplementary Figure 1: Distribution of TL across different measurement platforms.** TopWGS TelSeq TL (raw) indicates raw telomere length estimates derived from TelSeq in (Kbp) across 482,848 participants (see methods), qPCR (adjusted) TL indicates T/S adjusted values released by *Codd et al* across 462,675.

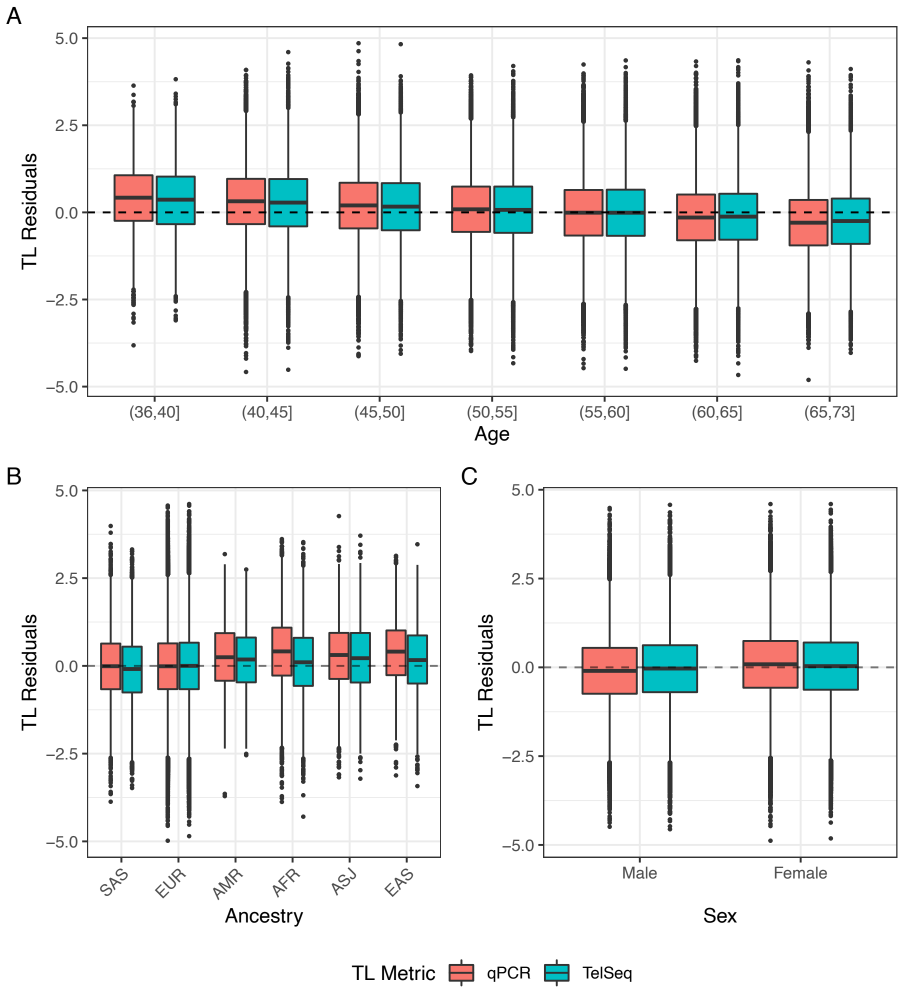


**Supplementary Figure 2: Age, Ancestry, and sex relationships with TelSeq TL measurements**. For each panel y axes denote telomere lengths residuals after regressing out age, sex, or ancestry depending on the x axis variable. **(A)** Boxplot of age by telomere residuals, Note that for age, each additional year was associated with an average loss of 10bp (+-0.2bp, P~0) in telomere length - an estimated supported by TopMed(Taub et al. 2022). **(B)** As above but for ancestral group (SAS = South Asian, EUR = Non-Finnish Europeans, AMR=Hispanic, AFR=African, ASJ=Ashkenazi Jewish and EAS=East Asian. Using EUR as baseline, SAS age and sex corrected WGS TL estimates were significantly shorter (-42bp +/- 9bp, P=1.7 x $10^{-21}$) whereas AFR (49bp + - 9bp, P=7.6 x $10^{-24}$) EAS (77bp +- 17bp, P=3.5 x $10^{-18}$) and AMR (79bp +- 31bp, P=4.5 x $10^{-7}$) were longer. **(C)** Females had significantly longer TL (28bp +-2.5bp, P=3.5 x $10^{-106}$) than males, after adjusting for age and ancestry.

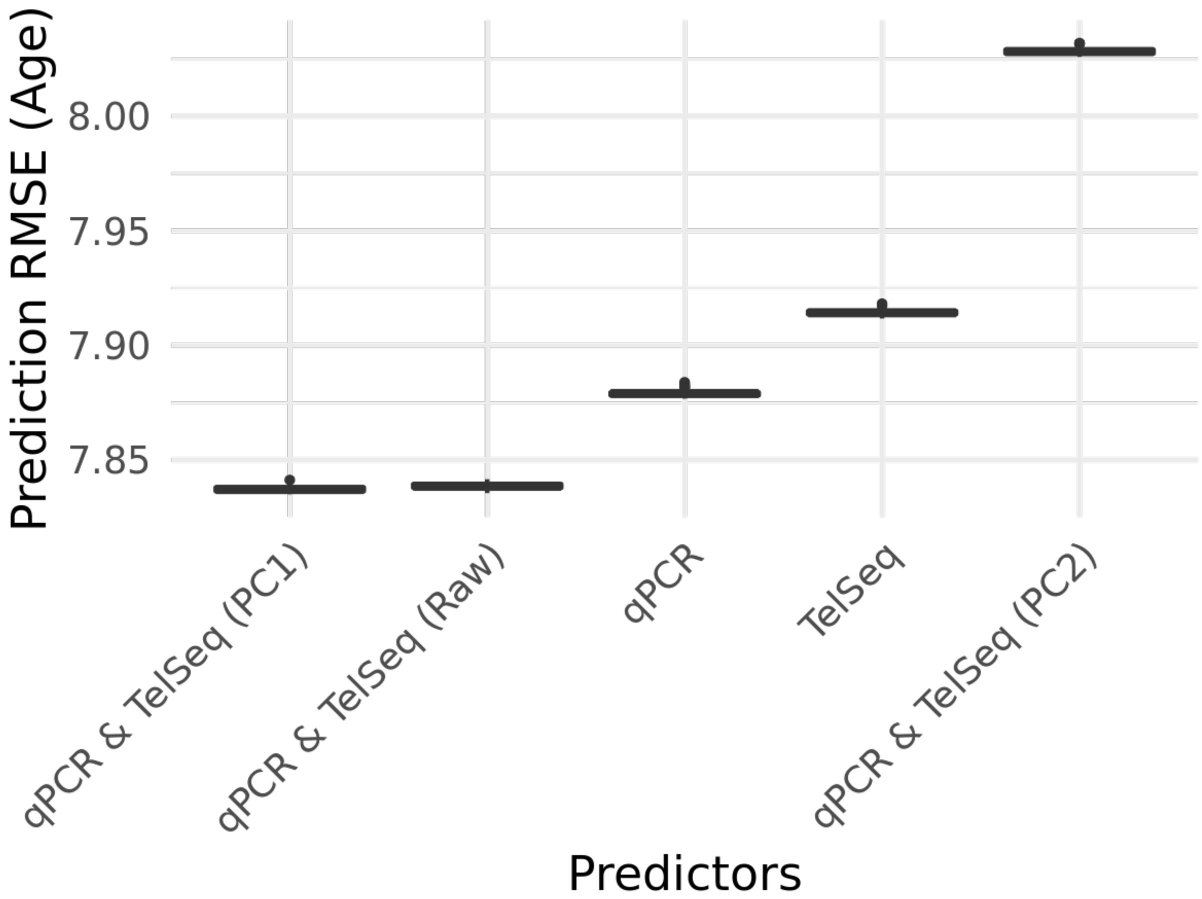


**Supplementary Figure 3**: **Box plot of age prediction for different TL metrics**. qPCR & TelSeq (PC1/PC2) indicates PCA derived composite score while qPCR & TelSeq (Raw) indicates the joint model (i.e age ~ $TL_{qPCR}$ + $TL_{TelSeq}$). Y axis indicates root mean squared error (RMSE) of age prediction using a training set of 10,000 samples and applying the resultant model to the remaining held out samples, distributions were derived from carrying out this procedure 100 times (see methods).

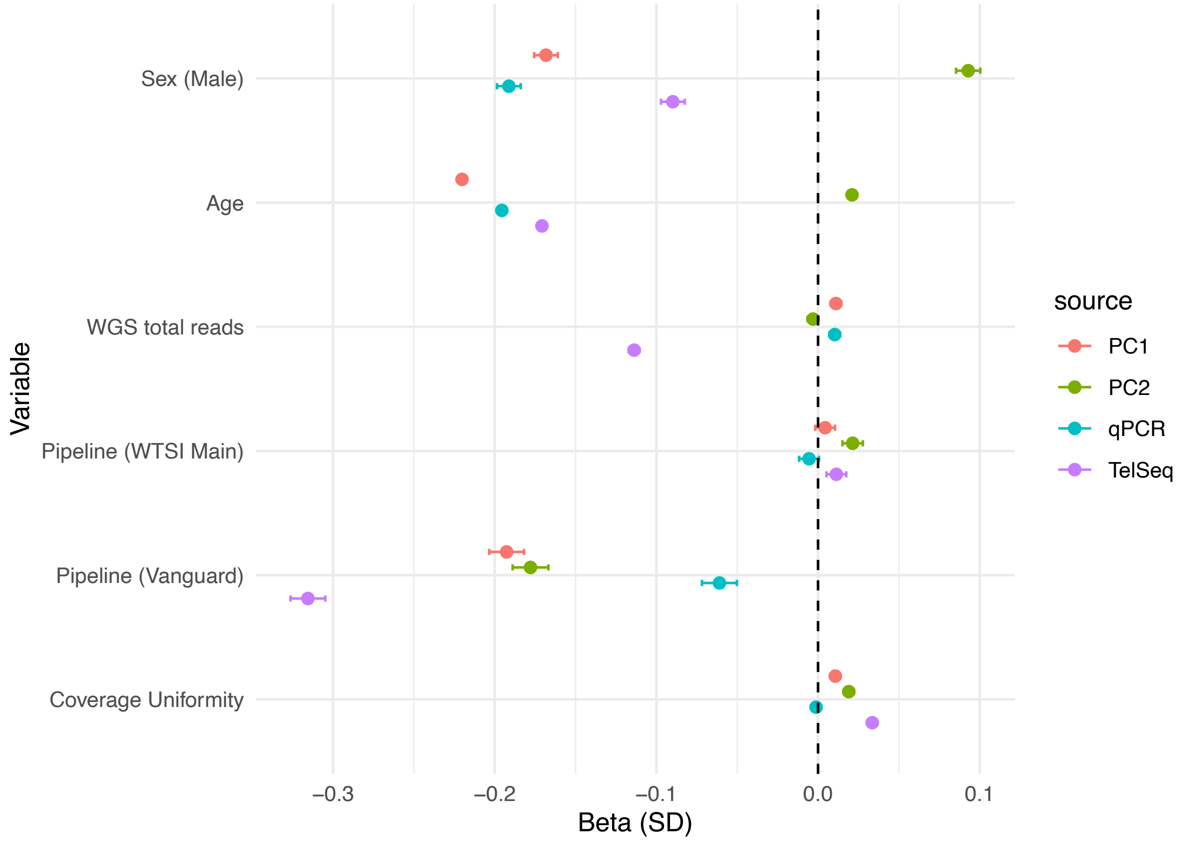


**Supplementary Figure 4: Multivariate analysis across different WGS measurement metrics.** We fit a linear model to combined (PC1 and PC2) as well as individual QC metrics including various WGS QC metrics as well as age and sex as positive controls. Pipeline metrics are with respect to the DeCODE WGS pipeline. A full table of results is available in **supplementary table 3**.

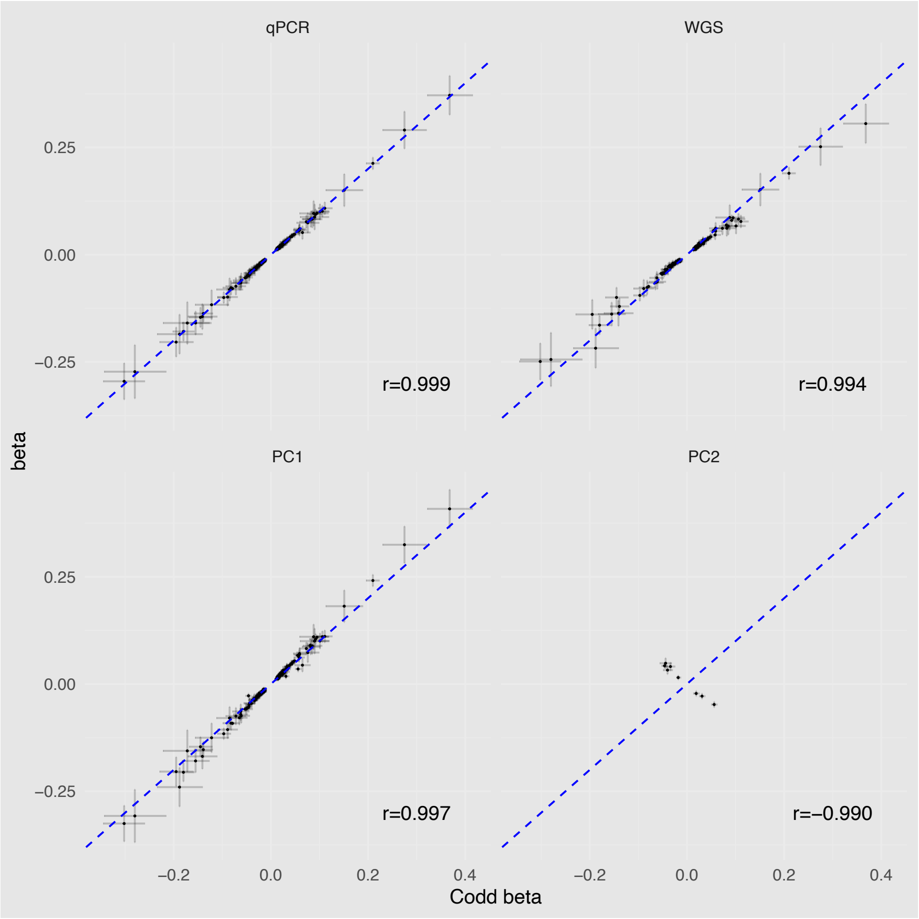


**Supplementary Figure 5: Comparison of GWAS effect sizes with Codd et al.** (y axis) for different TL measurements effect sizes with EUR only effect sizes from Codd et al (x axis). Crosses indicate 95% confidence intervals; Pearson's correlation coefficient are labelled on each panel.

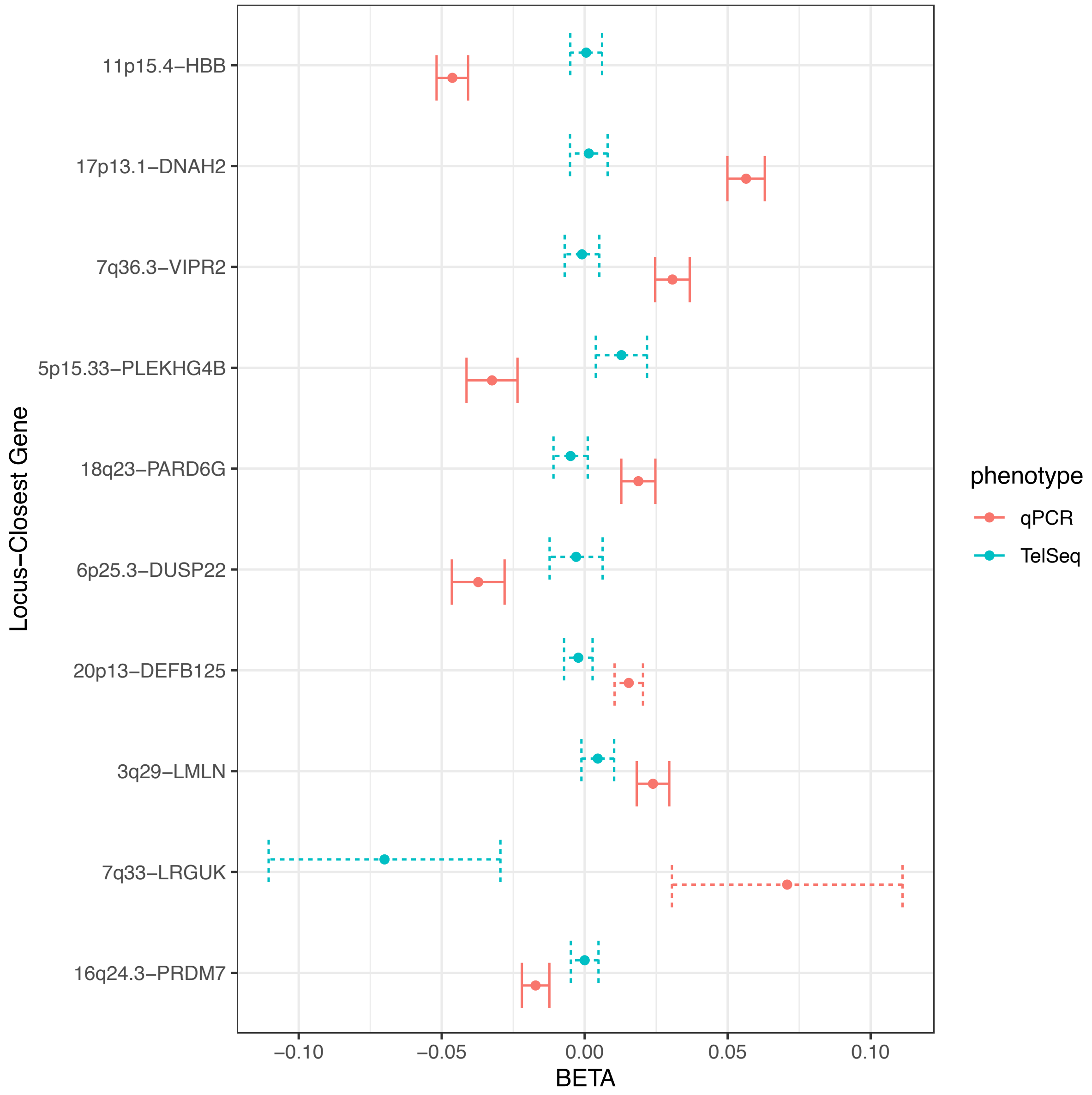


**Supplementary Figure 6: Forest plot of qPCR and TelSeq association statistics for significant PC2 loci.** Dotted error bars (95 % confidence intervals) indicate that the association was not genome-wide significant ($p<5 \times 10^{-8}$). Y axis is ordered by PC2 significance. PC2 is driven mainly by associations with qPCR that are not found to be associated with TelSeq, however it does highlight two loci where there is evidence of opposing effects at 5p15.33 and 7q33.

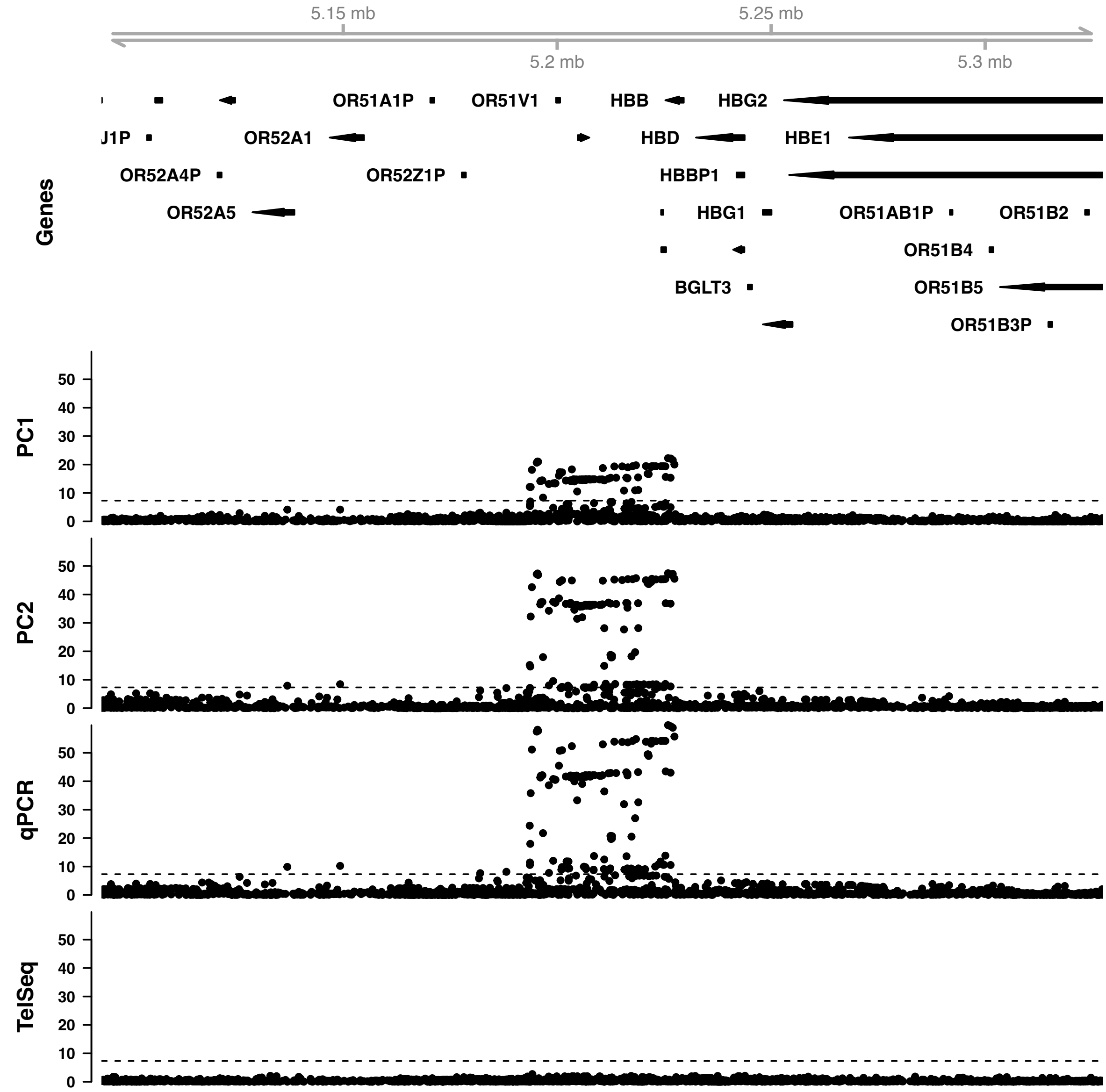


**Supplementary Figure 7: Comparison of GWAS summary statistics for *HBB* association.** Coordinates are for GRCh38, stanza show -log10(p) for different TL metrics., dotted line indicates p 5 x $10^{-8}$

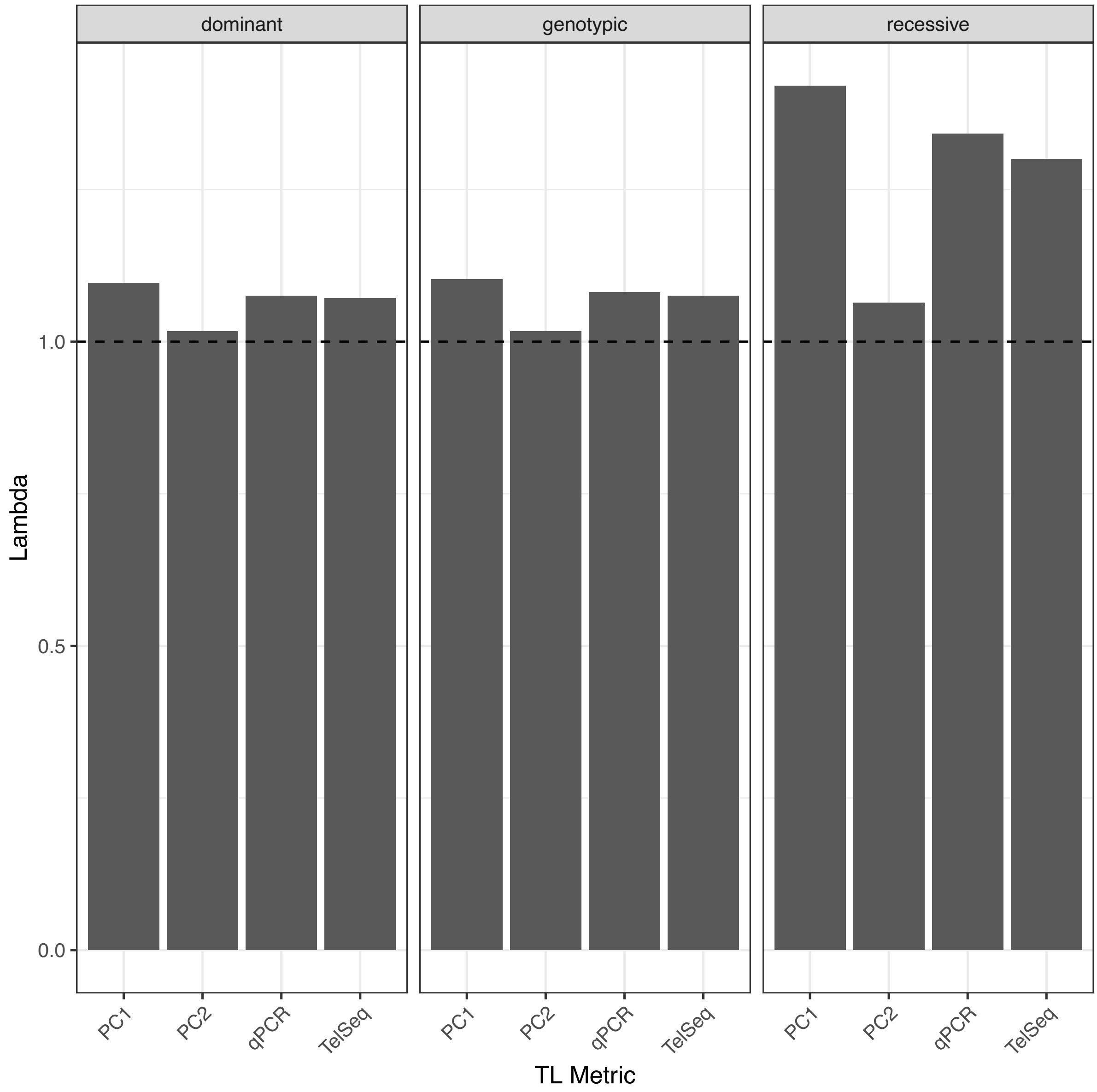


**Supplementary Figure 8: Genomic control across ExWAS models.** Y-axis indicates lambda genomic control showing that generally inflation is well controlled for genotypic and dominant models. There is some evidence for inflation in the recessive model however no significant associations were reported for this model.

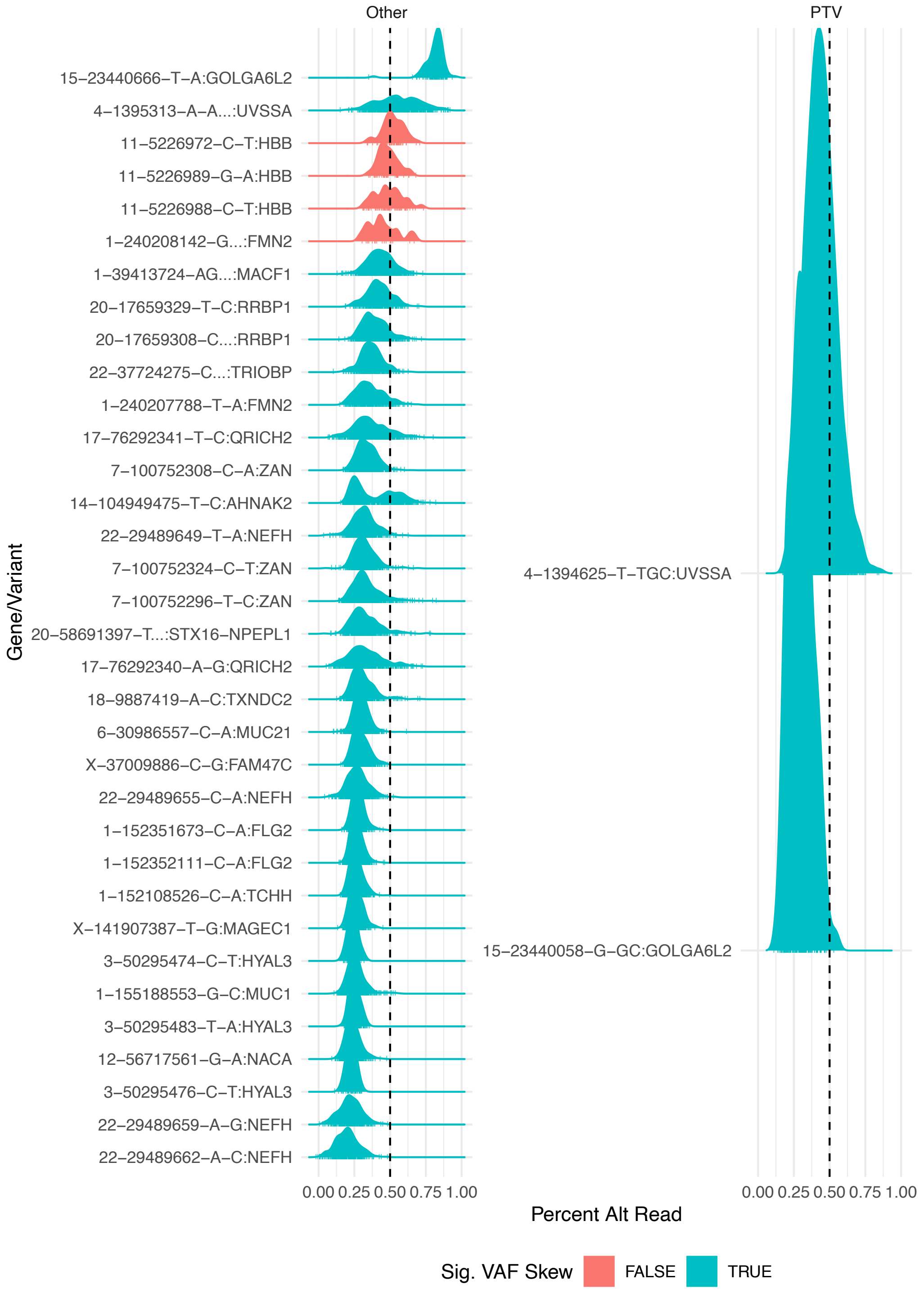


**Supplementary Figure 9: Density rug ridge plot of percentage of alternative allele reads for heterozygous individuals for PC1 ExWAS associations that also overlap a PC2 ExWAS associations**. Sig variant allele fraction (VAF) skew indicates variants that when combined across individuals show significant departure from expected ratio of 1:1 (horizontal dotted line) for alt and ref reads assessed by a binomial test. The rug ends (lines) indicate the percentage of alternative allele reads for heterozygous of individuals. As can be seen precluding *HBB* and *FMN2* loci all other variants show evidence for read imbalance, indicating that PC2 is indeed detecting spurious associations.

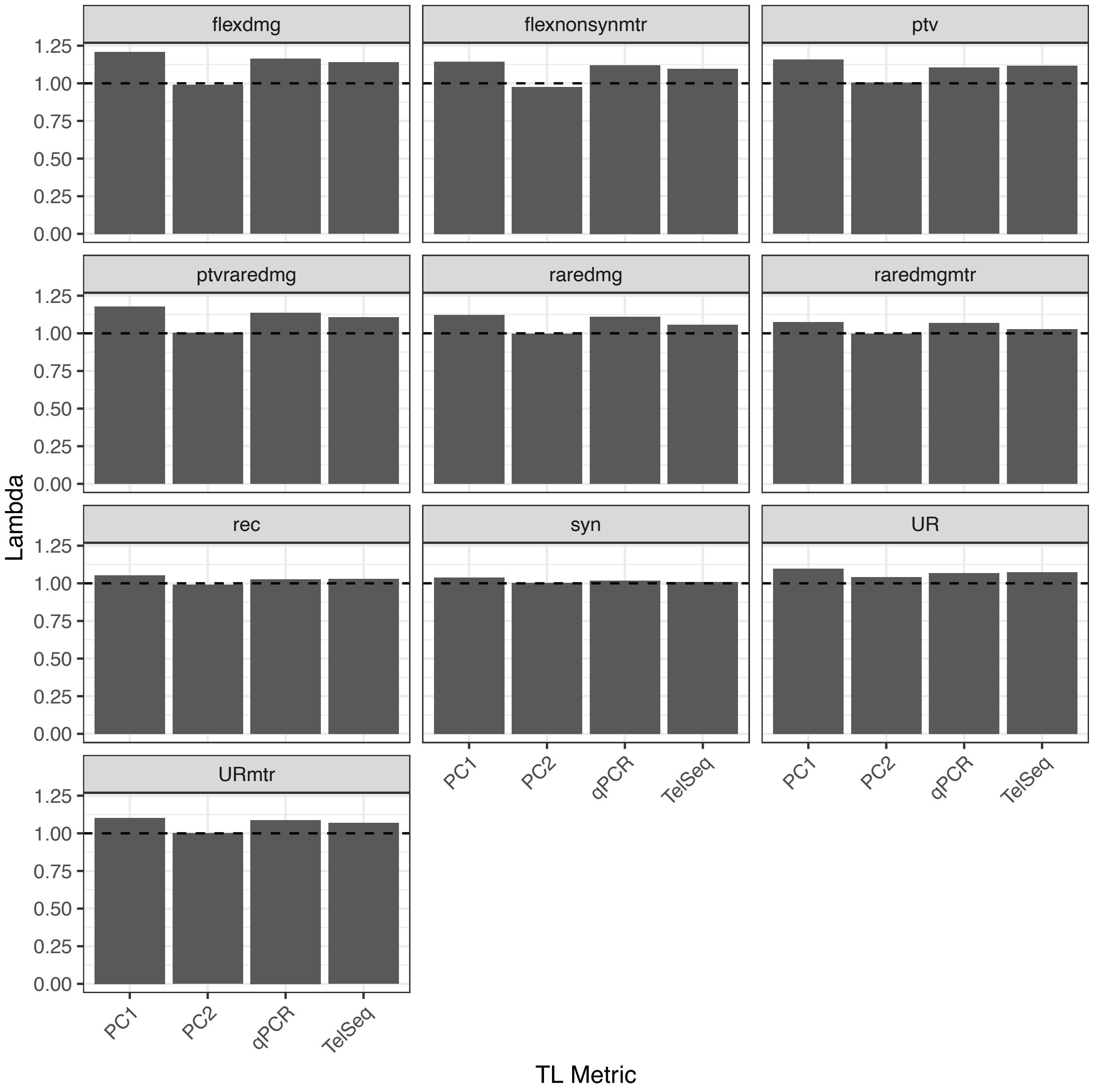


**Supplementary Figure 10: Genomic control across 10 qualifying variant collapsing models.** Y-axis indicates lambda genomic control showing that inflation is well controlled across all models.

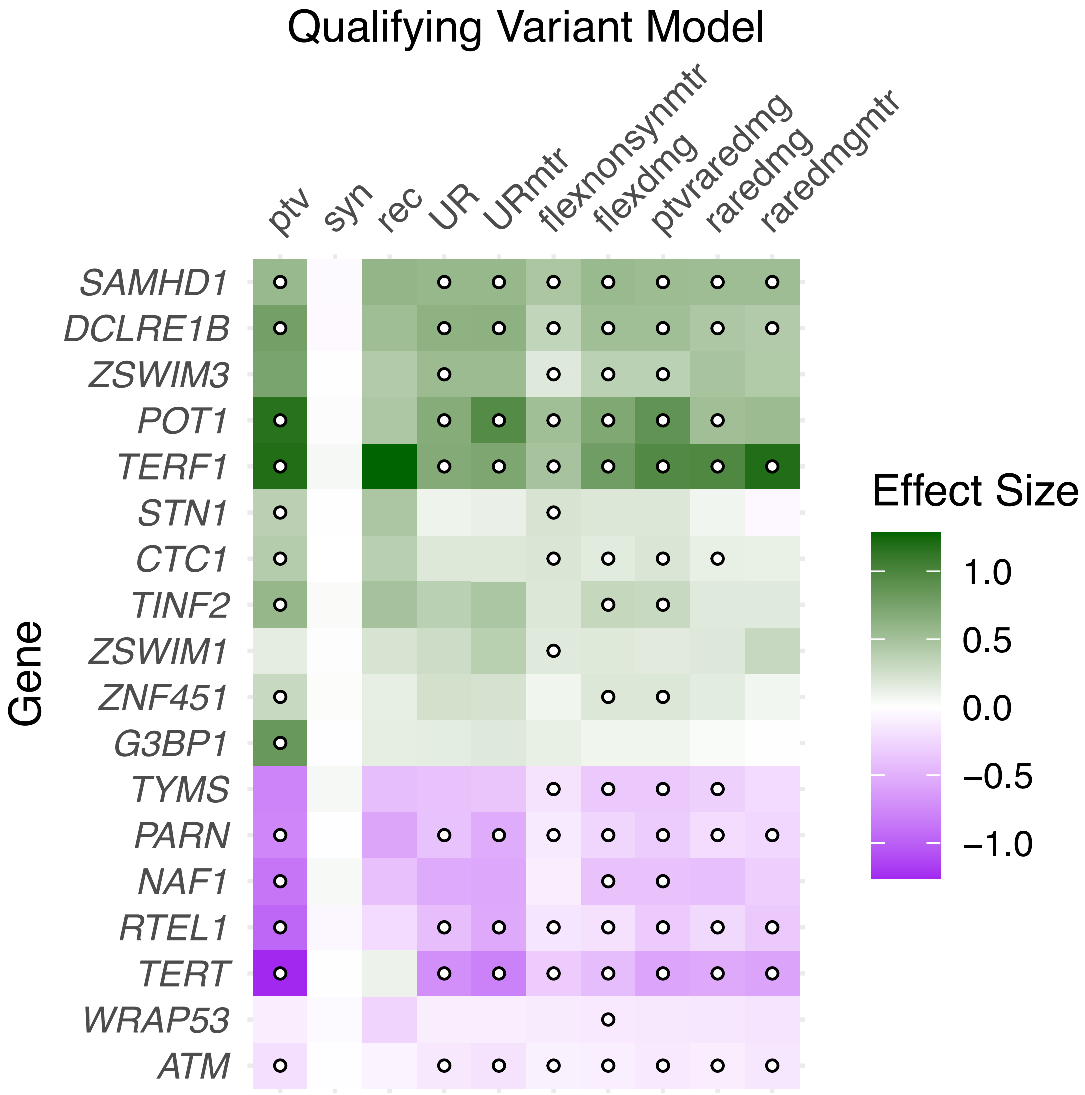


**Supplementary Figure 11: Heatmap of genome-wide significant genes from a gene collapsing analysis**. Shading indicates effect size (green = increased TL, purple = decreased TL), points indicate genome-wide significance (p≤1x10-8). The x-axis indicates the different qualifying variant models implemented which are described fully in Wang et al.. Briefly, *ptv*= rare protein truncating variants, *UR* = ultra rare variants, *URmtr* = ultra rare variants in missense intolerant regions (MTR), *raredmg* = rare damaging (REVEL) variants, *raredmgmtr* = as raredmg but with additional MTR filter, *flexdmg* = flexible non-synonymous, *flexnonsynmtr* = as flexdmg but with additional MRT filter, *ptvraredmg* = Union of ptv and raredmg models, *rec* = recessive model, *syn* = synonymous variants (negative control).

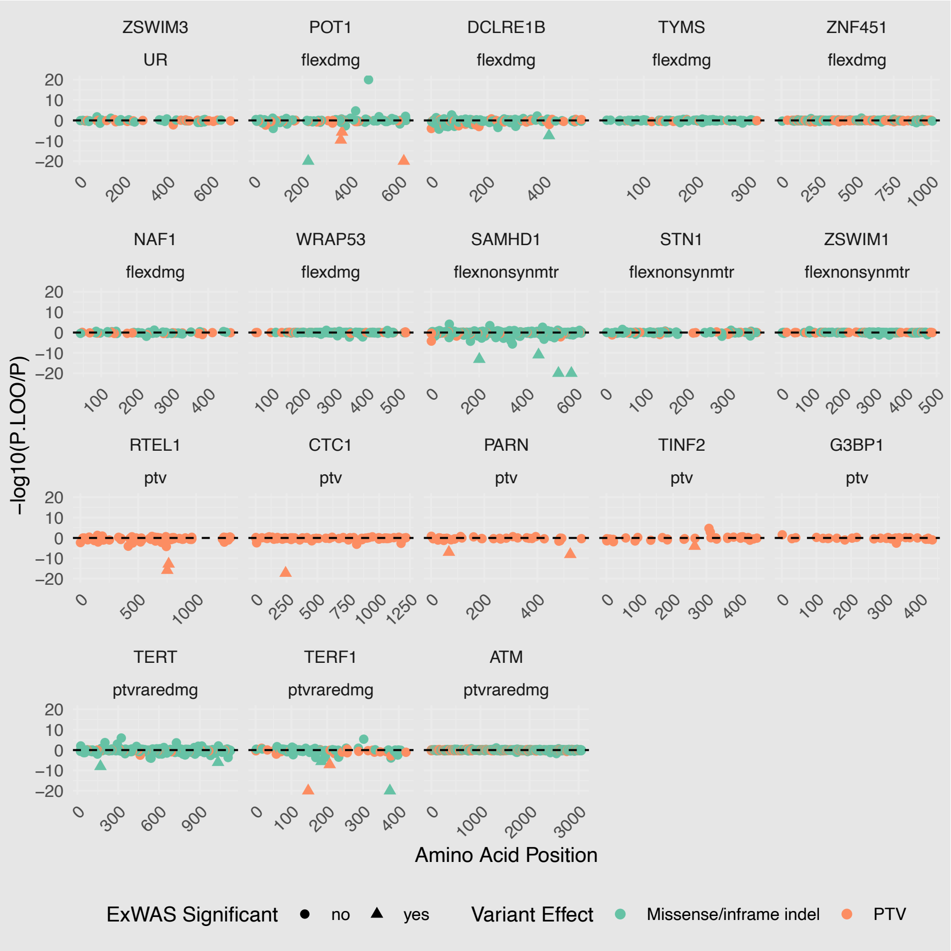


**Supplementary Figure 12: Leave-one-out collapsing analysis of significant genes for most significant qualifying variant model**. Points indicate variant left out and are coloured by function. Amino acid positions are based on canonical transcripts, triangular points indicate variants also found to be significant in ExWAS analyses. For clarity axes are truncated at -20 and 20.

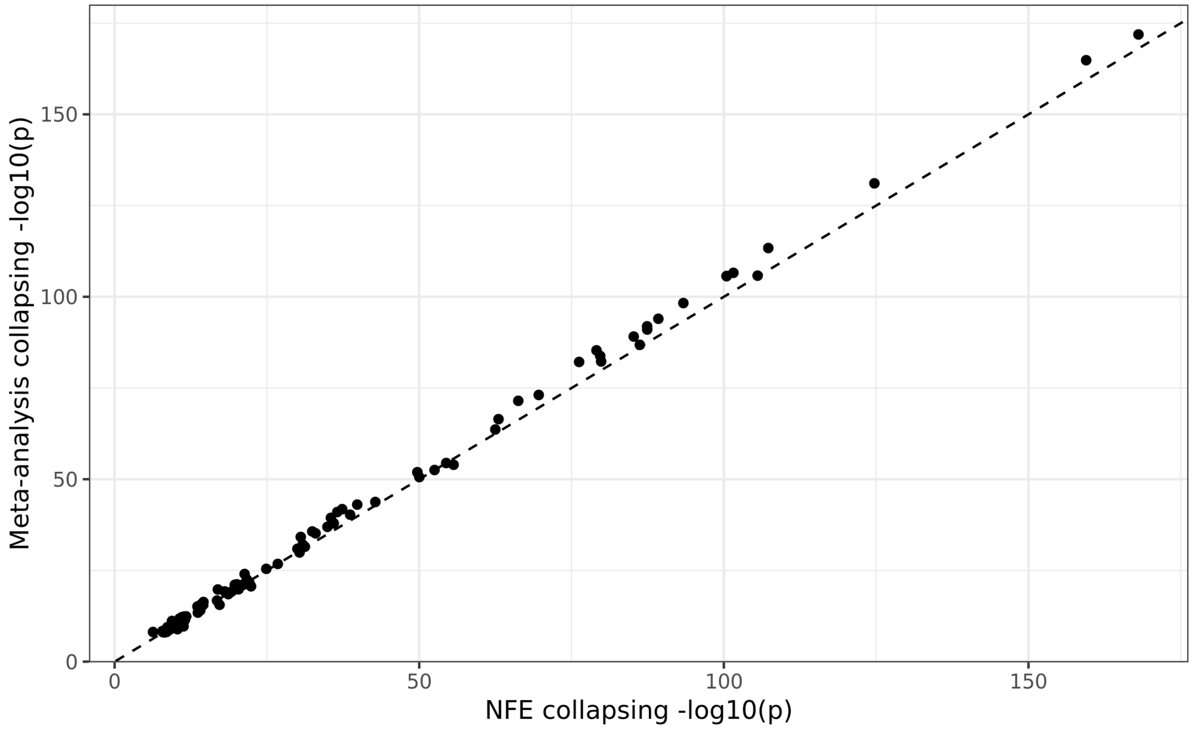


**Supplementary Figure 13: Comparison of p values for NFE and fixed effect cross ancestry meta-analysis collapsing analysis (NFE,AFR,SAS,EAS, and ASJ).** Only variants $p_{NFE}$<5 x $10^{-5}$ are shown.

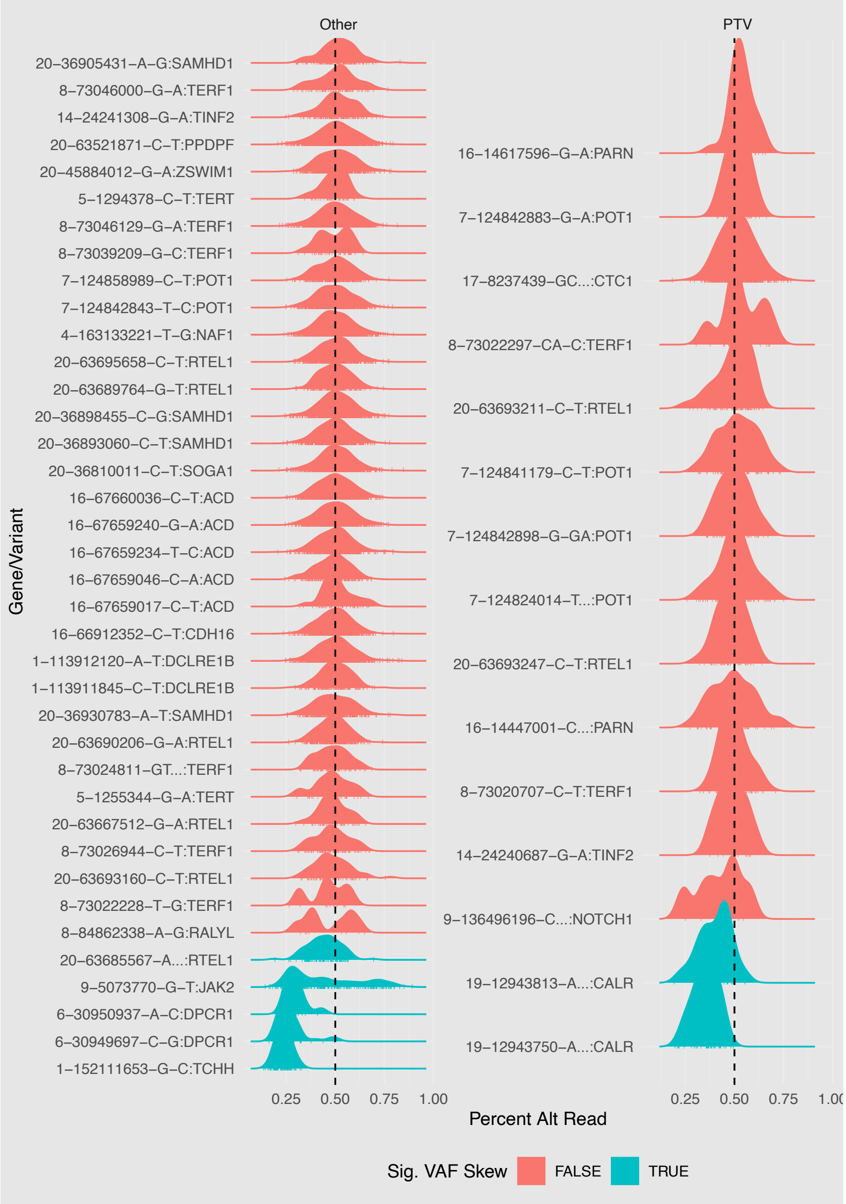


**Supplementary Figure 14: Density rug ridge plot of % Alt Reads (i.e the fraction of reads corresponding to the alternative read) for heterozygous individuals.** Sig VAF Skew indicates variants that when combined across individuals show significant departure from expected ratio of 1:1 (horizontal dotted line) for Alt and Ref reads assessed by a binomial test. The rug ends (lines) indicate the % Alt Read for heterozygous of individuals.

**Supplementary Note on GWAS comparison with *Codd et al.***

We compared our GWAS results with a previously published GWAS that used qPCR TL measurements on the same set of participants. As expected, effect sizes from our GWAS on qPCR, WGS, PC1 and PC2 TL were all highly correlated with their qPCR-based effect sizes (**Supplementary Figure 5**) and we were able to replicate associations reported as significant by *Codd et al.* ($p<8.3x10^{-9}$, N Loci=131) at 131, 81, 124 and 7 loci, across the respective TL measures (**Supplementary Table 5**).
